# Long-term health-related quality of life in pediatric ECMO survivors: a prospective controlled study

**DOI:** 10.1101/2025.10.08.25337633

**Authors:** Alexiane Le Hellaye, Oscar Werner, Nicolas Joram, B Gaillard Le Roux, Amanda Guerra, Olivier Cadeau, A Chenouard, Arnaud Roy, Alban-Elouen Baruteau, A Gaultier, Beatrice Desnous, Pascal Amedro, Pierre Bourgoin, the QUALIREHAB study group

## Abstract

**Introduction:** Extracorporeal Membrane Oxygenation (ECMO) is an intervention used for neonates, infants, and children with severe respiratory or cardiac failure. While ECMO has been linked to improved survival rates, it is also associated with neurological complications and potential long-term effects on health-related quality of life (HRQoL), when compared to healthy peers. In order to better characterize the longer-term outcomes of ECMO itself, this study evaluated HRQoL, as well as neurodevelopmental, motor, and cognitive outcomes in pediatric ECMO survivors in comparison to a matched control group.

**Methods:** We conducted a prospective cross-sectional matched case-control study at Nantes University Hospital, France. Forty-two ECMO survivors supported with ECMO from January 2014 to January 2023 were matched with 42 pediatric intensive care unit (PICU) survivors. HRQoL was assessed using the PedsQL 4.0 Generic Core Questionnaire. Neurodevelopmental outcomes were evaluated with the Age and Stage Questionnaire (ASQ) or the Strengths and Difficulties Questionnaire (SDQ), motor function with the Global Motor Function Classification Score (GMFCS), and executive functioning with the Behavior Rating Inventory of Executive Function (BRIEF).

**Results:** Overall HRQoL scores were slightly lower in the ECMO group, with a mean adjusted difference for HRQoL total score of -5.4, 95%CI -10.6 to -0.3, *p=0.040*. Notably, physical functioning was lower in the ECMO group, with a mean adjusted difference of -9.6, 95%CI -16.8 to -2.5, *p=0.009*. Amongother outcome measures reported, executive function complaints affected approximately one thirds of the cohort and were associated with impaired HRQoL. Among other variables tested, parental stress index and parental level of education were also associated with HRQoL total score or subdomains.

**Conclusion:** ECMO survivors often experience slight changes in their health-related quality of life as compared with a matched control cohort, particularly in physical functioning. This study also identifies cognitive and executive functioning impairments that may persist long after discharge from the PICU.

## Introduction

Neonates, infants and children admitted to pediatric intensive care units (PICUs) with severe respiratory or cardiac failure can benefit from extracorporeal membrane oxygenation (ECMO). Survival rates are approximately 54% when ECMO is used for heart failure, 61% for pediatric respiratory conditions, and up to 73% for neonatal respiratory distress ^1^. Among survivors, nearly half present with moderately or severely abnormal neurodevelopmental status at hospital discharge ^2^. Such impairment is probably explained by the high incidence of brain injury during ECMO support, with 20 to 40% of patients experiencing acute neurological events such as clinical or electrical seizures, or ischemic or hemorrhagic strokes ^3–6^. Risk factors for neurological complications include age (with younger children at higher risk due to immaturity of the blood-brain barrier), underlying condition (higher risk for heart failure indications), and occurrence of cardiac arrest preceding or occurring at the time of ECMO cannulation ^5,7^.

The impact of ECMO support on long-term health-related quality of life (HRQoL) is a significant concern for both clinicians and families ^8^. Multiple studies have described long-term HRQoL after ECMO support in specific populations, such as congenital diaphragmatic hernia, cardiac disease, cardiac arrest, or in mixed ECMO cohorts ^9^. These studies consistently reported similar patterns of altered neurodevelopmental outcomes in ECMO survivors.

Additionally, with the recent insights from post-intensive care evaluations, it has become evident that some severely ill children may also experience long-term HRQoL impairment ^10^, making it challenging to assess the specific impact of ECMO on long-term outcomes. Most studies exploring late neurodevelopmental outcomes, functional status, or HRQoL after ECMO have reported results as standardized differences compared with healthy controls, with the exception of a few studies conducted in ECMO survivors cannulated for congenital diaphragmatic hernia ^11,12^ or after congenital heart surgery ^13^. From these studies, the impact of ECMO itself on long term outcomes appeared limited, as these specific populations are known to present long-term neurodevelopmental impairment independent of the severity of initial presentation ^14,15^.

The objectives of this study were to evaluate HRQoL in pediatric ECMO survivors compared with matched controls, and to identify factors associated with impaired HRQoL.

## Materials and Methods

### Study design and ethics

We carried out a comparative prospective cross-sectional matched case-control study at Nantes University Hospital, Nantes, France, a regional referral hospital for pediatric ECMO and pediatric cardiac surgery ^16^. An independent national review board approved the study (CPP Est III: 2023-A00262-43). This study was registered in the clinical trials database (NCT05721105). Written parental consent and/or child consent was obtained before evaluation.

### Study population

Patients treated with ECMO in Nantes University Hospital within a ten-year period (January 2014-January 2023) were eligible. Patients were contacted using both mailed correspondence and telephone communication. We included survivors aged greater than 2 years old and less than 18 years at the time of study inclusion. The exclusion criteria included: known genetic disorders or congenital malformation syndromes that could lead to severe neurodevelopmental deficits, parental refusal to participate in the study, and language barriers that prevented complete informed consent.

A matched comparative group was selected from patients hospitalized during the same study period. Controls were selected if they received mechanical ventilation for at least 48 hours, vasoactive support, and/or extracorporeal renal replacement therapy. Matching criteria included: age (neonate and infant under 3 months, infant 3-24 months, children 2-5 y, children 5-10 y), year of hospital stay, diagnostic category (respiratory failure, cardiac failure, or cardiopulmonary resuscitation), and diagnostic sub-category (congenital diaphragmatic hernia, congenital heart disease (CHD), post-cardiac surgery low cardiac output syndrome, or septic shock).When more than one control case was identified, the first patient approached was selected by hospital admission order. Patient characteristics and outcomes were recorded from a local database. Data describing ECMO support (type and mode of support, complications) were prospectively recorded in the ELSO database (Nantes University Hospital, ELSO center number 389).

### Primary outcome: health-related quality of life (HRQoL)

HRQoL was assessed with PedsQL 4.0 Generic Core Questionnaire^17^. Parent proxy reports were performed for all children, and self-reports for children ≥ 5 years, as recommended^18^. This validated instrument applies to various ages and health conditions and evaluates HRQoL across multidimensional scales (physical functioning, 8 items; emotional functioning, 5 items; social functioning, 5 items; school functioning, 5 items) and 3 summary scores (total scale score; physical health summary score, 8 items; and psychosocial health summary score, 15 items). Over the last month, each item was evaluated according to a 5-point response scale: 0, never a problem; 1, almost never a problem; 2, sometimes a problem; 3, often a problem; and 4, almost always a problem. A simplified 3-point response scale was used for the self-reports of younger children. Mean scores were calculated for each dimension, resulting in a score ranging from 0 to 100, higher values indicating better HRQoL.

### Secondary outcomes

The Ages and Stages Questionnaire (ASQ-3), completed by parents, was used to evaluate neurodevelopmental (NDV) outcomes in children aged 2-5 y ^19,20^ . NDV delays were detected by comparing individual scores with the age-specific cut-off values defined in the ASQ-3 manual, corresponding to 2 standard deviations below the normative mean for each domain. Five domains were assessed: communication, fine motor, global motor, problem-solving, and social interaction. Children were classified into 3 categories according to ASQ scores: OPTIMAL (all domains above cut-off), INTERMEDIATE (one domain below cut-off), and NON-OPTIMAL (two or more domains below cut-off).

The Strengths and Difficulties Questionnaire (SDQ), completed by parents, was used to evaluate behavioral and emotional outcomes in children aged 6-18 y ^21^ . SDQ consists of a 25-item questionnaire focusing on the behavioral sphere (emotional symptoms, conduct problems, hyperactivity/ inattention, peer relationship problems, prosocial behavior). The total score allowed classification of patients into three categories: 0-15, normal; 16-17, borderline; > 17, abnormal.

The Gross Motor Function Classification System (GMFCS) was used to assess global motor disability in all age categories. This parent-completed tool has demonstrated good inter-rater reliability between parents and healthcare professionals ^22^ . The GMFCS is a 5-level classification: level 1 indicates no motor disability, level 2 mild disability, and levels 3-5 were grouped as severe motor disability. The Behavior Rating Inventory of Executive Functions (BRIEF-2) and the Preschool version (BRIEFP), for patients under 5 years old, were used to assess executive function disorders. These validated questionnaires have been widely used in pediatric population, including chronically ill children with CHD and various other chronic conditions ^23–26^ . The BRIEF is an 86-item Likert-scale inventory that evaluates the frequency (never, sometimes, often) of difficulties in everyday executive functioning. It comprises eight clinical scales: inhibit, shift, emotional control, initiate, working memory, plan/organize, task monitor, and organization of materials. Age- and sex-specific norms were used to convert raw scores into T-scores. A cut-off score defined as 1.5 standard deviations above the mean (i.e., T ≥ 65), consistent with the manual, was applied to indicate clinical significance.

The Parental Stress Index (PSI) was completed by parents to assess parental stress. The PSI is a validated 36 items questionnaire ^27^ . A raw score > 90 indicated a high level of parental stress. The respondent was either the father or the mother, and responses were considered as proxy reports. Parental educational level was recorded for both parents, but only maternal educational educational level was used for analysis, classified as high school degree or not.

In addition, information on patients’ living arrangements, schooling, and specialized medical or rehabilitation follow-up was collected to provide an overview of long-term support needs.

### Statistical analysis

Continuous data are presented as medians with interquartile range [IQR] or means ± standard deviation, and categorical data as numbers (n) and percentages (%). Non-parametric Mann–Whitney U tests or parametric Student’s t-test were used for continuous variables, χ2 test for categorical variables, and Kruskal-Wallis test for ordinal variables in intergroup comparisons (ECMO group versus matched control group). Paired tests were applied to compare different scores withing the same patient. Pearson correlation analysis was also performed. Variables associated with HRQoL were explored using ANOVA (analysis of variance). Multivariate analysis was conducted with ANCOVA (analysis of covariance). Stepwise inclusion was applied to select informative variables, based on the Akaike Information Criterion.

## Results

### Participants

A total of 155 patients underwent ECMO support in our center between January 2014 and December 2023 and 76 (49%) survived. Among survivors, 62 (82%) were eligible for the study, and 42 (67.7%) completed the questionnaires and were included in the analysis (**Figure 1**, flowchart). Each ECMO survivor was successfully matched with a control according to predefined criteria. For reference, detailed descriptions of ECMO patients and their matched controls are provided in **Supplemental Material (Table S1)**.

**Figure 1:**
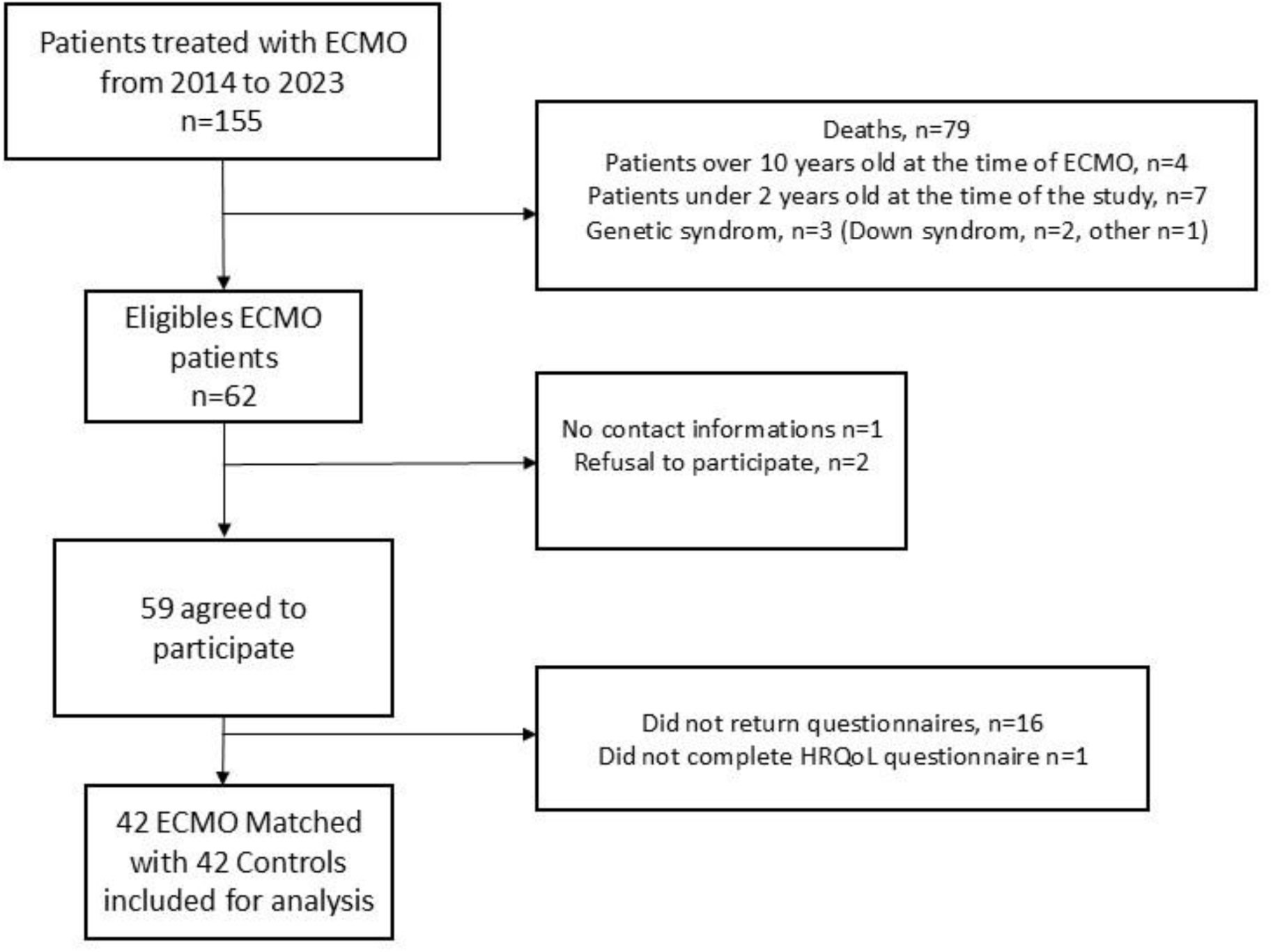
Flowchart.

The mean age at inclusion was 7.4±3.6 years, and the mean time elapsed between hospital admission and inclusion was 5.4±2.4 years. A summary of patient characteristics and outcome measures for both groups is presented in **Table 1**. Characteristics of ECMO patients are detailed in **Table 2**. The median age at ECMO initiation was 100.5 days [IQR 2.8-1622], with nearly half (19/42, 45.2%) supported during early infancy (< 3 months). Most patients (32/42, 83.3%) received venoarterial ECMO, with cervical cannulation in the majority of cases (21/35; 60%). Four patients (9.3%) experienced an acute neurological event (ANE) during ECMO.

**Table 1.** Summary of patients included in the ECMO group (n=42) and matched control group (n=42)

|  | ECMO group | Matched control group | *Sig.* |
| --- | --- | --- | --- |
| Age Category |  |  |  |
| • Neonates/infants 0-3 months | 20 (47.62%) | 18 (42.86%) | - |
| • Infants 3-24 months | 9 (21.43%) | 10 (23.81%) |  |
| • Children 2-5 yo | 4 (9.52%) | 7 (16.67%) |  |
| • Children 5-10 yo | 9 (21.43%) | 7 (16.67%) |  |
| Age at admission (days, median [IQR]) | 100.5 [3.5,1495.5] | 143.5 [4.5, 1495] | 0.859 |
| Gender (M/F) | 21 (50%)/21 (50%) | 26 (61.9%)/16 (38.1%) | 0.379 |
| MV time (days), Mean (SD) | 10.69 (± 9.15) | 6.63 (± 9.91) | 0.056 |
| LOS, ICU (days) | 21.67 (± 19.33) | 12.36 (± 16.93) | **0.021** |
| LOS, Hospital (days) | 51.93 (± 35.51) | 34.48 (± 43.42) | **0.047** |
| Age at inclusion |  |  |  |
| • 2-4 y | 8 (19.05%) | 9 (21.43%) | - |
| • 5-7 y | 13 (32.50%) | 12 (28.57%) |  |
| • 8-12 y | 19 (47.50%) | 17 (40.48%) |  |
| • 13-18 y | 2 (4.76%) | 4 (9.52%) |  |
| Age at inclusion (years) | 7.3 (±3.4) | 7.6 (±3.9) | 0.721 |
| Time elapsed between admission and survey (years) | 5.7 (±2.44) | 6.0 (±2.5) | 0.567 |
| Questionnaire Filled |  |  |  |
| • PedsQL, proxy form | 42 | 42 | - |
| • PedsQL, Self reported | 34 | 33 |  |
| • ASQ 24m, 36m, 48m or 60m | 12 | 13 |  |
| • SDQ | 30 | 30 |  |
| • GMFCS | 42 | 41 |  |
| • PSI | 42 | 42 |  |
| • BRIEF (preschool) | 13 | 14 |  |
| • BRIEF | 29 | 28 |  |

**Table 2.** Summary of characteristics of patients (n=42) supported with ECMO (defined according to ELSO recommendations).

|  |  |
| --- | --- |
| ECMO support type (n, %) |  |
| • Cardiac | 25 (59.5%) |
| • Respiratory | 13 (30.9%) |
| • ECPR | 4 (9.5%) |
| Initial ECMO mode (n, %) |  |
| • VA, cervical | 21 (50%) |
| • VA, central | 14 (33.3%) |
| • VA, femoral | 2 (4.8%) |
| • VV, double lumen | 5 (11.9%) |
| Run par patient (n, %) |  |
| • 1 run | 42 (100%) |
| • > 1run | 0 |
| ECMO duration (days) mean±SD | 5.7±4.1 days |
| Complications (n, %) |  |
| • Any complication (ELSO definition) | 17 (40.4%) |
| • ANE (ELSO definition) | 4 (9.3%) |

ANE: Acute Neurologic Event (Seizure, stroke, Transient or persistent neurologic deficit). ECPR: Extracorporeal cardiopulmonary resuscitation. ELSO: Extracorporeal life support organization. VA: Veno arterial. VV: veno venous.

### Primary outcome: HRQoL

All proxy-reported HRQoL questionnaires were completed (n=84), and self-reports were available for all children aged ≥ 5 years: 34/34 (100%) in the ECMO and 33/33 (100%) in controls. Overall, mean total HRQoL scores were similar between children and their parents: 74.2±16.3 versus 74.3±17.4, respectively (*p=0.201)*. In the 67 parent-child pairs, scores were highly correlated (r = 0.93, 95% CI [0.88, 0.96], *p < 0.001*). Between groups comparisons showed no significant differences: in proxy-reported total HRQoL, 71.4±17.3 vs. 77.2±17.3 (ECMO vs controls, *p=0.132*); in self-reported total HRQoL, 70.9±15.3 vs. 77.7±16.8 (ECMO vs controls, *p=0.088*); Details of HRQoL subdomains are reported in **Table 3**, and **Figure 2** provides a graphical summary. Individual paired values are shown in **Supplemental Figure S1.**

**Figure 2:**
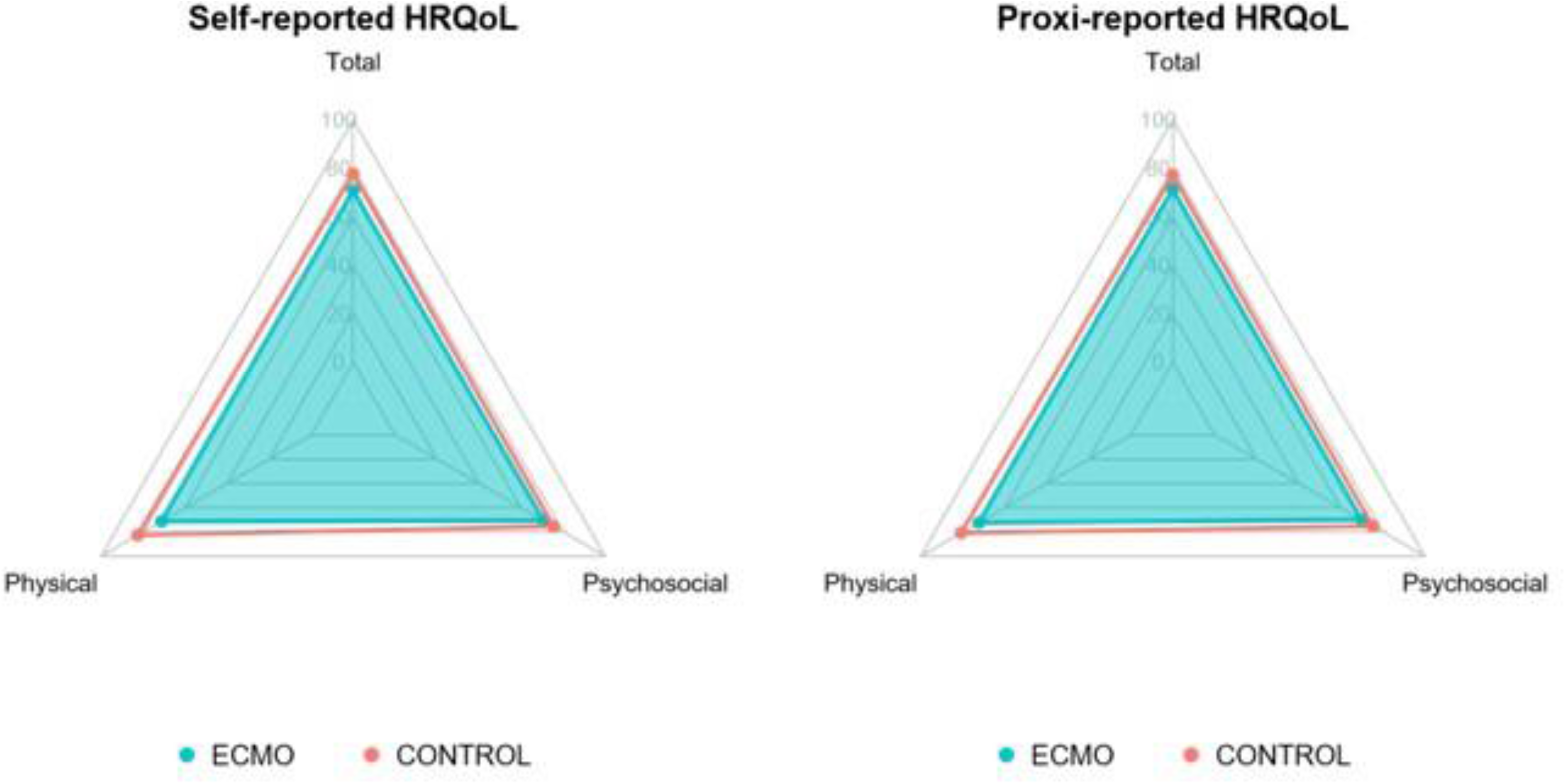
Left panel: Self-reported HRQoL. ECMO group (blue line), Matched group (red line). Right panel: Proxi-reported HRQoL. ECMO group (blue line), Matched group (red line).

**Table 3.**
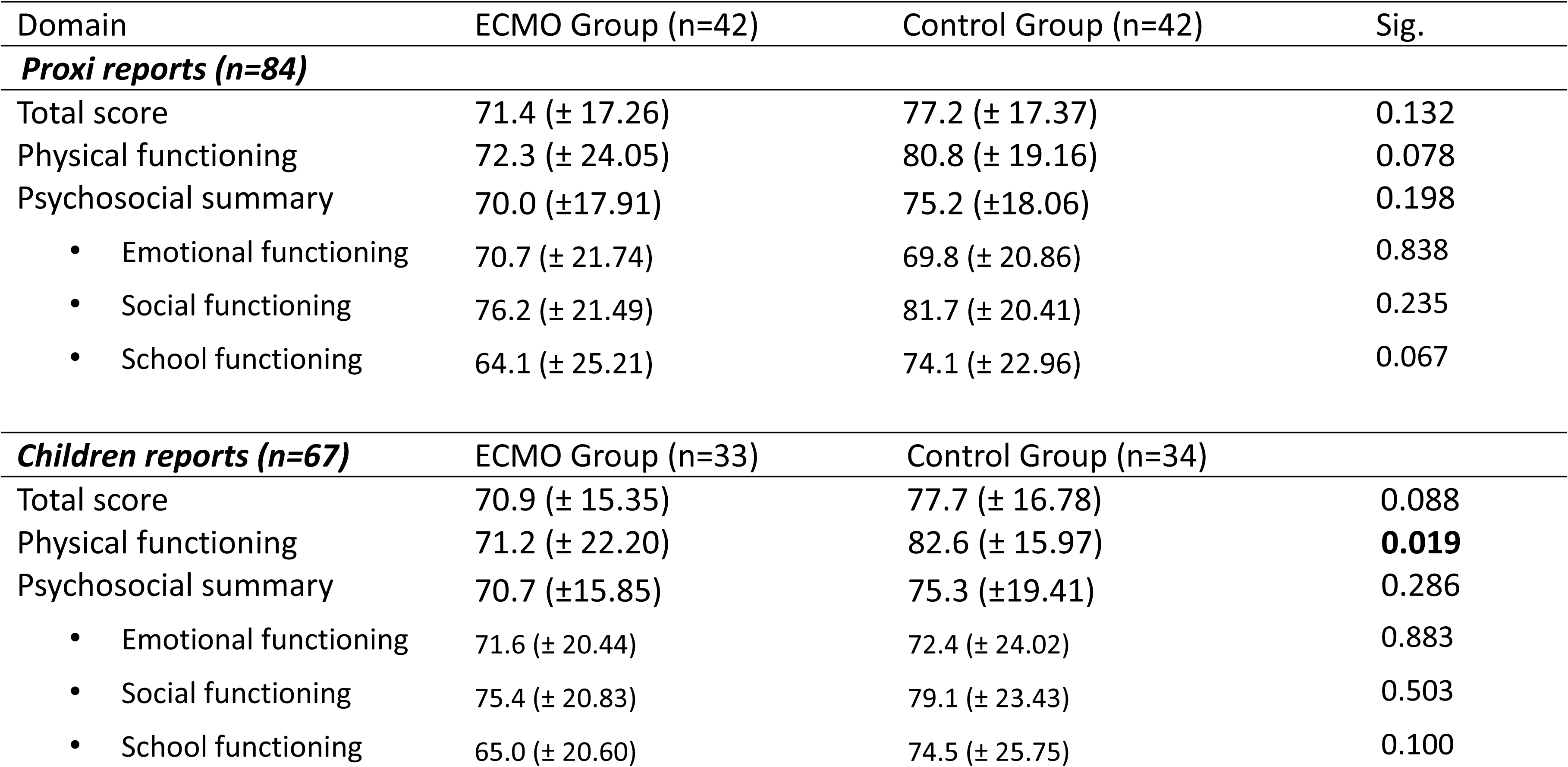
Summary of HRQoL evaluation in the ECMO group and matched Control Group.

### Factors associated with HRQoL

**Table 4** summarizes the results of the multivariate analysis, while the corresponding univariate analyses are provided in **Supplemental Table S2**. In multivariate analysis, the independent predictors of lower total HRQoL scores were: ECMO support (−5.4, 95% CI −10.6 to −0.3, p = 0.040), extracorporeal cardiopulmonary resuscitation (ECPR) (−9.7, 95% CI −18.7 to −0.7, p = 0.035), high parental stress (PSI >90: −14.9, 95% CI −22.5 to −7.3, p < 0.001), and executive function disorders (BRIEF T ≥65: −17.4, 95% CI −24.5 to −11.4, p < 0.001). A graphical representation of adjusted mean differences between ECMO and control groups is shown in **Figure 3**, illustrating the negative impact of ECMO on total and physical HRQoL, but not on psychosocial HRQoL.

**Figure 3:**
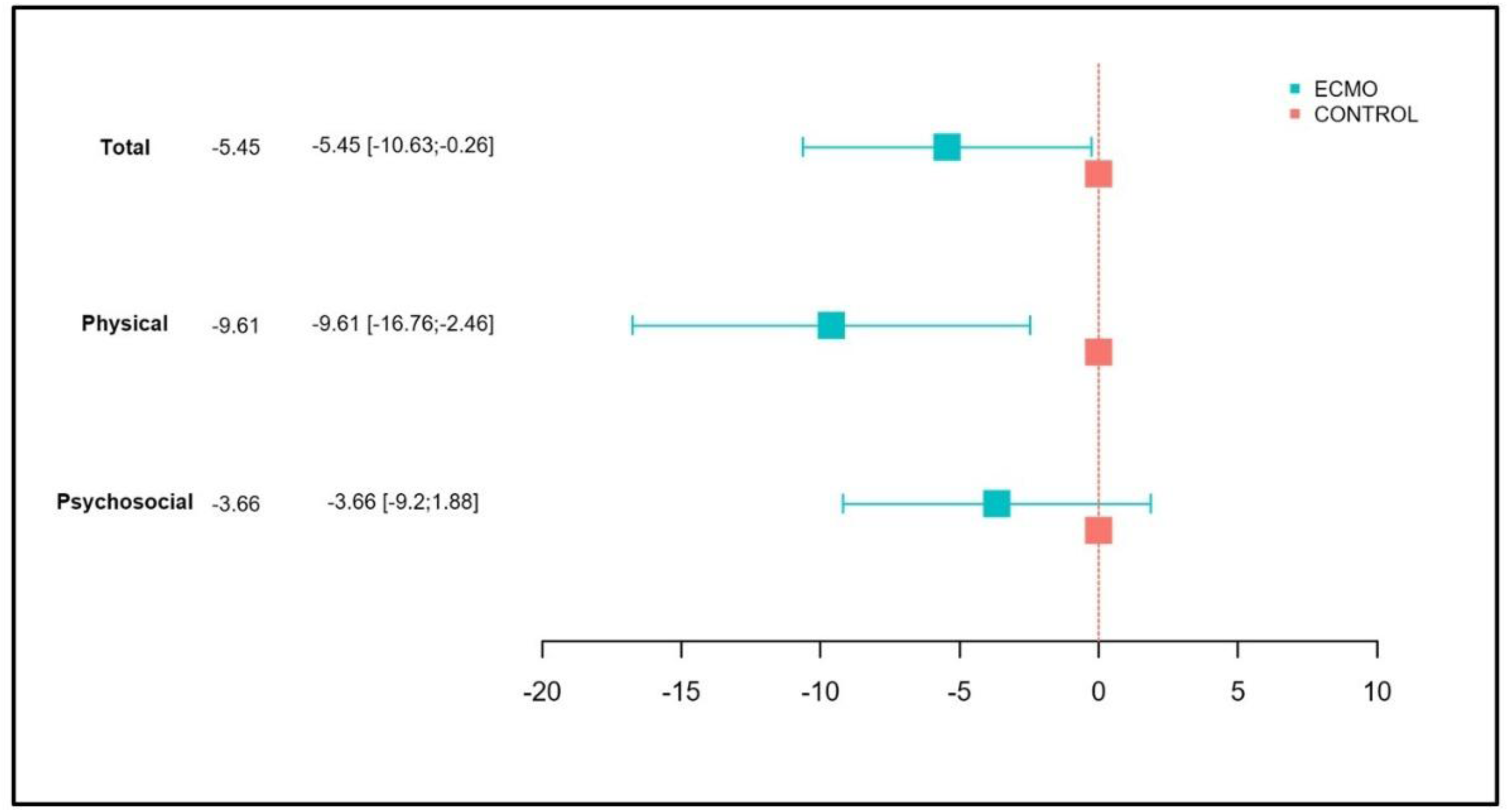
Forrest plot representation of multivariate linear regression for HRQoL total score and components.

**Table 4.** Multivariate analysis for HRQoL total score, physical and psychosocial components (proxi reports). Values are reported for variables selected after stepwise inclusion according to Akaike Information Criterion only. Bold letters indicates p value < 0.05.

|  | HRQoL total score |  |  | Physical functioning |  |  | Psychosocial functioning |  |  |
| --- | --- | --- | --- | --- | --- | --- | --- | --- | --- |
| variable | Estimate | aCI95% | P value | Estimate | CI95% | P value | Estimate | CI95% | P value (multi) |
| **Age at admission** |  |  |  |  |  |  |  |  |  |
| CHILDREN 6-10Y | ref |  |  | ref |  |  |  |  |  |
| NEONATE 0-3 mo | 3.87 | [-3.3;11.0] | 0.283 | 13.3 | [0.2; 26.5] | **0.047** |  |  |  |
| INFANTS 3-24 mo | -5.94 | [-14.0;2.1] | 0.147 | -1.9 | [-14.7; 10.9] | 0.764 |  |  |  |
| CHILDREN 2-5Y | -0.72 | [-10.1;8.6] | 0.878 | 3.4 | [-9.9 ; 16.7] | 0.610 |  |  |  |
| **Age at inclusion** |  |  |  |  |  |  |  |  |  |
| 2-4 years |  |  |  | Ref |  |  | Ref |  |  |
| 4-7 years |  |  |  | -1.7 | [-12.3; 8.9] | 0.753 | -18.9 | [-30.1 ; -7.8] | **0.001** |
| 7-12 years |  |  |  | 8.8 | [-2.6; 20.3] | 0.126 | -18.6 | [-29.7 ; -7.5] | **0.001** |
| 12-20 years |  |  |  | -4.4 | [-24.1; 15.2] | 0.655 | -29.5 | [-46.8 ; -12.1] | **0.001** |
| **Sexe** |  |  |  |  |  |  |  |  |  |
| F |  |  |  |  |  |  | Ref |  |  |
| M |  |  |  |  |  |  | -7.6 | [-15.6 ; 0.3] | 0.060 |
| **Status** |  |  |  |  |  |  |  |  |  |
| Matched controls | ref |  |  | ref |  |  | ref |  |  |
| ECMO | -5.4 | [-10.6;-0.3] | **0.040** | -9.6 | [-16.8; -2.5] | **0.009** | 0.934 | [-7.3 ; 9.1] | 0.821 |
| **Diagnostic category** |  |  |  |  |  |  |  |  |  |
| Cardiac | ref |  |  | Ref |  |  |  |  |  |
| ECPR (Cardiac Arrest) | -9.69 | [-18.7;-0.7] | **0.035** | -25.2 | [-38.0; -12.3] | **< 0.001** |  |  |  |
| Respiratory | -4.28 | [-10.2;1.7] | 0.157 | -3.6 | [-12.3; 5.1] | 0.408 |  |  |  |
| **LOS (hospital)** |  |  |  |  |  |  |  |  |  |
| 0-30 days |  |  |  |  |  |  | Ref |  |  |
| 30-60 days |  |  |  |  |  |  | -15.1 | [-24.5 ; -5.8] | **0.002** |
| > 60 days |  |  |  |  |  |  | -9.2 | [-19.9 ; 1.3] | 0.087 |
| **Parental stress Index** |  |  |  |  |  |  |  |  |  |
| Raw score < 90 | ref |  |  | Ref |  |  |  |  |  |
| Raw score > 90 | -14.9 | [-22.5;-7.3] | **< 0.001** | -21.8 | [-32.9; -10.8] | **<0.001** |  |  |  |
| **Grade of education** |  |  |  |  |  |  |  |  |  |
| High |  |  |  |  |  |  | Ref |  |  |
| Low |  |  |  |  |  |  | -14.1 | [-23.3 ; -4.8] | **0.003** |
| **Executive Function disorders** |  |  |  |  |  |  |  |  |  |
| Stdd T-score < 65 | ref |  |  | Ref |  |  | Ref |  |  |
| Stdd T-score > 65 | -17.4 | [-24.5;-11.4] | **< 0.001** | -11.8 | [-20.8; -2.9] | **0.010** | -27.8 | [-36.2 ; -19.5] | **<0.001** |

For the physical functioning sub score, the independent predictors of lower scores were ECMO support -9.6, 95%CI -16.8 to 2.5, p=0.009), ECPR (-25.2, 95%CI -38.0 to -12.3, p< 0.001), high parental stress (-21.8, 95%CI -32.9 to -10.8, p<0.001)and executive dysfunction (-11.8, 95%CI -20.8 to -2.9], p=0.010), whereas younger age at inclusion was a predictor of better outcomes compared with older children (13.3, 95%CI 0.2 to 26.5, p=0.047).

For the psychosocial functioning sub domain, the independent predictors of lower scores were older age at inclusion (2-4 years at inclusion being the referral value), prolonged hospital stay (- 15.1, 95%CI -24.5 to -5.8, p=0.002), low maternal educational level (-14.1, 95%CI -23.3 to -4.8), p=0.003), and executive dysfunction (-27.8, 95%CI -36.2 to -19.5, p<0.001.

### Overview of patient’s needs

All interviewed patients reported living with their families, except for one ECMO survivor placed in foster care for parental reasons. All children older than 3 years at the time of inclusion were enrolled in school: 46 (66.6%) attended without assistance, 14 (20.3%) received support from a special needs assistant, and 9 (13%) followed another form of adapted schooling, with no significant difference between groups. Follow-up by a rehabilitation physician, speech therapist, psychologist or occupational therapist was needed in 8/84 (9.5%), 23/84 (27.4%), 9/84 (10.7%), and 11/84 (13.1%) patients, respectively, with no significant between-group differences.

### Other outcome measures collected

NDV was assessed with ASQ-3 in 25/25 (100%) of the patients aged ≤5 years at the time of inclusion, with a mean age 3.4±1.3 years and after 3.5±1.4 years from hospital admission. Overall, NDV was quoted optimal in 16/25 (64%), intermediate in 3/25 (12%) and non-optimal in 6/25 (24%), with 5 of them belonging to the ECMO group, this result being non-significantly different from the control group (*p=0.310*). SDQ was completed in 58/59 (98.3%) of the patients aged ≥ 6 years old at the time of inclusion, with a mean of 9.2±2.8 years and after 6.4±2.1 years after hospital admission. Behavioral and emotional outcome assessed through SDQ was normal in 40/58 (68.8%), borderline in 6/58 (10.3%), and abnormal in 12/59 (20.7%) of the patients, with an identical repartition between ECMO and control groups (21 vs 19; 3 vs 3 and 6 vs 6 patients were classified as having normal, borderline or abnormal behavioral and emotional SDQ score, *p=0.863*). Global motor functioning assessed with GMFCS was available in 83/84 (98.8%) of the patients included. No GMF impairment was noted in most of the patients included (72/83 (86.6%)), mild GMF impairment was noted in 7/83 (8.4%), including 6 patients belonging to the ECMO group, whereas a severe GMF was found in 4 (4.8%) of the patients, with 3 patients belonging to the ECMO group, conferring a difference between groups (*p=0.029*). All patients completed online BRIEF or BRIEF-P questionnaires according to their age at inclusion (**table 2**). Overall, 25/84(29.8%) of the patients analyzed depicted complaints about executive functioning (standardized T-score ≥ 65), with no difference between groups (*p=0.633*).

## Discussion

Our findings reinforce that, beyond survival, ECMO is associated with measurable impairments in quality of life. However, the fact that the overall quality of life between ECMO group and controls are different with a small gap (estimated adjusted difference: -5.4 [-10.2; -0.2]) is encouraging, as it suggests that despite the intensive nature of ECMO support, ECMO survivors can achieve comparable long-term quality of life outcomes compared with selected PICU survivors. This is consistent with the recent study by Michel et al., which reported that the quality of life in ECMO survivors is acceptable and comparable to patients with chronic illnesses ^28^. The difference observed for overall HRQoL must be analyzed in regard to studies reporting the minimal clinically important difference (MCID, expressed as a difference for HRQoL score), a metric that has been shown to vary across population studied, periods of time, age population, and pathologies. Among reports available, MCID was set at 6.1, 11, 7 to 11, or 4.5, in the context of children with diabetis, chronic rheumatologic illness, cerebral palsy, or 4 to 5 years after cardiac open heart surgery, respectively^29–32^.

This study further underscores the significant influence of impaired physical functioning on health-related quality of life (HRQoL), indicating that factors such as medical severity, for instance the need for ECMO support and cardiac arrest are primary contributors to physical limitations. Notably, the ECMO group exhibited considerable deficits in physical functioning, consistent with findings from previous studies^13,33,34^ . Long-term concerns have been reported in children with a history of critical illness, regardless of documented neurological injury during PICU stay. In Ducharme et al., about 40% of children assessed two years post-PICU showed fine or gross motor dysfunction^10^. Interestingly, better outcomes among younger patients may reflect greater neuroplasticity and recovery potential early in life. This suggests early rehabilitation after PICU discharge could improve HRQoL, as seen recently in adolescents and young adults post-congenital heart surgery^35^.

Another significant finding from the measured outcomes was that a considerable proportion of patients or their proxies reported executive function concerns, as assessed by the BRIEF questionnaire, in approximately one-third of both the ECMO patient group and the matched control group, with no significant differences observed. Although cognitive impairments have been documented in children with congenital heart disease [38], this study is, to the best of our knowledge, the first to identify such deficits among the most critically ill PICU patients^36^. In this study, psychosocial functioning was primarily linked to parental stress, maternal education, and executive dysfunction, with no significant association observed with severity of illness. These results indicate that cognitive impairment may be related to the broader effects of a PICU stay, which are influenced by illness severity and the family and social environment, rather than being directly caused by ECMO. Previous research has documented the effects of parental stress on children’s outcomes^37,38^. It is important to recognize these impacts and consider providing support for parents, such as access to psychological resources in addition to medical follow-up, to help improve long-term outcomes for children.

Strengths and limitations. To our knowledge, this is the first study including matched PICU controls specifically designed to analyze the long-term impact of ECMO on quality of life. Our cohort of 84 patients, some of whom were followed up to 10 years post-PICU discharge, offers a valuable longitudinal perspective on both QoL and the potential for recovery over time. A major strength of this study is the high response rate (71.2%) and the comprehensive evaluations conducted, which included both cognitive and executive function assessments, providing a robust picture of the patients’ long-term outcomes. Additionally, the matched control group of critically ill PICU patients enhances the validity of our comparisons by accounting for the severity of illness in both groups and allows us to better isolate the specific impact of ECMO versus critical illness alone.

However, several limitations must be acknowledged. First, this is a single-center study with a relatively small sample size, which may limit the generalizability of our findings to other institutions. Moreover, the small number of patients with acute neurological events (ANE) restricts the power of our subgroup analysis. Our analysis was based on data from the ELSO registry, and we did not conduct a detailed analysis of brain imaging reports, as systematic brain imaging is not part of routine care in ECMO survivors in our unit. Recent studies ^39^ have demonstrated the relationship between white matter lesions on post-operative MRI studies and neurodevelopmental outcomes (NDV) following cardiac surgery under cardiopulmonary bypass. A detailed neuroimaging assessment could provide further insight into the potential underlying mechanisms of cognitive impairment. Additionally, while our study assessed parental and child-reported HRQoL, we did not collect data from both parents, which could provide a more nuanced understanding of the family’s overall perception of the child’s health and well-being. If patients were included after various time periods without any observed impact from the time elapsed since PICU admission on HRQoL, there remains a lack for a longitudinal perspective. It must be acknowledge that different trajectories are observed after PICU admission, with some patients recovering to baseline HRQoL 6 months after PICU admission ^40^. Given that patients in this study were examined around 5 years after PICU admission, it is probable that these findings reflect long-term outcomes, although the ultimate trajectory of patients remains uncertain. Parental stress is reported here, and it is known that posttraumatic stress syndrome (PTSS) may affect children with a history of severe illness as part of the post intensive care syndrome (PICS)^40^. Whether PTSS influences health-related quality of life (HRQoL) in children after severe critical illness has not been reported, its evaluation could be valuable for this specific population.

In conclusion, this study offers valuable insights into the long-term health-related quality of life of pediatric ECMO survivors, revealing both areas of resilience and specific domains of vulnerability. While overall HRQoL seems lower, with uncertained identification of a clinically important difference as compared to matched PICU controls, impairments in physical functioning were particularly evident. This study underscores the potential critical role of early neurological monitoring and rehabilitation in mitigating deficits. Future research should focus on targeted interventions for both patients and their families to enhance recovery and quality of life after ECMO support.

## Data Availability

Data available on request to

## Acknowledgements

PB thanks CIC-FEA, Nantes University Hospital, and all ECMO program participants— perfusionists, surgeons, and PICU staff—for their support.

